# Accuracy and completeness of death recording in primary care electronic health records compared with national death registrations in England using OpenSAFELY-TPP

**DOI:** 10.64898/2026.10.14.26365028

**Authors:** Martina Pesce, William J Hulme, Irene Kyomuhangi, Em Prestige, David Evans, Siu Ying Wong, Ben Goldacre, Sebastian CJ Bacon, Amir Mehrkar, OpenSAFELY Collaborative, Brian MacKenna, Andrea L Schaffer

## Abstract

**Introduction:** Primary care electronic health records (EHRs) are an important resource for health research, but death recording in primary care EHRs may be incomplete or differ from Office for National Statistics (ONS) death registrations. We used OpenSAFELY-TPP to assess the completeness and concordance of death recording and provide recommendations for using date-of-death information.

**Methods:** Using linked primary care EHRs and ONS death registrations, we assessed whether ONS-registered deaths from 2020–2025 had a primary care date of death and, where present, calculated the difference between dates. Where a primary care date was absent, we assessed death-related SNOMED CT codes. Analyses were conducted overall and by year, population subgroup, and general practice. We also examined GP registration end dates and primary-care-recorded deaths without a corresponding ONS record, including monthly patterns near the data extraction date.

**Results:** Among 1,409,275 ONS-registered deaths, 88% had the same date recorded in primary care and 8% had no corresponding primary care date. Among 1,324,995 deaths recorded in primary care, <2% had no corresponding ONS record. Deaths recorded in only one source were more frequent among people aged <45 years and those from Mixed, Asian or Asian British, Black or Black British, and Chinese or Other ethnic groups. ONS-only dates were also more frequent in London and the West Midlands. Primary-care-only deaths increased close to date of data extraction, reaching 7% in February 2026. Same-day GP deregistration increased from 9% in 2020 to 85% in 2025.

**Conclusions:** Death recording in primary care EHRs showed high completeness and concordance with ONS registrations, but discrepancies remained for recent deaths and some population groups. Data recency and population-specific completeness should therefore be considered when using linked mortality data for research.

## Introduction

Primary care electronic health records (EHRs) have become an important resource for health research, despite being collected primarily to support direct patient care and healthcare administration rather than research. Mortality is a key outcome in health research, with applications including monitoring population mortality trends^1^, evaluating the effectiveness and safety of medical products^2,3^, and quantifying health inequities^4–6^. As these analyses rely on the accurate ascertainment of deaths, understanding the completeness and quality of mortality data is essential for interpreting research findings and minimising the risk of bias.

In England, the Office for National Statistics (ONS) death registration database is considered the national reference source for mortality data^7^. Civil death registrations are based on statutory reporting processes and undergo multiple validation and quality assurance procedures involving different institutions^8^. The ONS death registration database is updated regularly with newly available or amended registrations^8^.

Mortality information can also be captured in primary care EHRs, including through dedicated date-of-death fields and clinical codes indicating death, recorded using SNOMED CT (Systematized Nomenclature of Medicine Clinical Terms), the standard terminology for clinical coding in NHS primary care^9^. However, there is no statutory requirement to record deaths in primary care EHRs, and practices may not always know when a registered patient dies, which may contribute to incomplete or delayed recording. As a result, some deaths may be missing from primary care EHRs, and recorded dates of death may not always correspond exactly to those in the ONS death registration database^10,11^.

Understanding the extent and patterns of these differences is important for research using primary care EHRs. If deaths are consistently under-recorded in specific populations, overall mortality estimates and comparisons between groups may be affected. GP registration status is also relevant, as registration end dates are frequently used to define study eligibility, follow-up, and censoring in epidemiological research. Understanding how GP registration end date relates to the date of death, including whether registration ends before, shortly after, or substantially after death, is therefore important for the interpretation and use of primary care EHRs in research. OpenSAFELY is a secure analytics platform that provides access to pseudonymised primary care EHRs from approximately 24 million patients registered with general practices using the TPP SystmOne software system, covering around 43% of general practices in England^12^. Using patient NHS numbers, these records are linked to additional healthcare and administrative datasets, including ONS death registrations from 2019 onwards^13^.

Using OpenSAFELY, we compared death information recorded in primary care EHRs with ONS death registrations. We specifically (1) assessed the completeness and concordance of primary care death records compared with ONS mortality records; (2) examined the temporal relationship between ONS mortality records’ dates of death and GP registration end dates; and (3) assessed the completeness of ONS mortality records compared with deaths recorded in primary care. Lastly, we provide recommendations for the use of date-of-death information in OpenSAFELY-TPP, considering both ONS mortality data and primary care records.

## Methods

### Study design and period

We conducted a descriptive observational study covering the period from 1 January 2020 to 28 February 2026. The start date corresponded to the first complete year for which both ONS and primary care data were available, and the end date to the last complete month before the ONS data import on 6 March 2026. The analyses used data available in OpenSAFELY on 15 May 2026. Comparisons between data sources were restricted to 2020–2025. Deaths occurring between 2025 and 2026 were analysed separately to assess the completeness of the ONS mortality register database compared with deaths recorded in primary care near the data extraction date.

### Data sources

All data were linked, stored and analysed securely using the OpenSAFELY platform, https://www.opensafely.org/, as part of the NHS England OpenSAFELY COVID-19 service. Data include pseudonymised data such as coded diagnoses, medications and physiological parameters. No free text data are included. No GP data from patients who have registered a Type-1 Opt-out with their GP surgery were included. Analysis was executed using R version 4.4.1 (2024-06-14 ucrt). All code is shared openly for review and re-use under MIT open license [https://github.com/opensafely/death-report/]. Detailed pseudonymised patient data is potentially re-identifiable and therefore not shared.

In OpenSAFELY, death-related information is sourced primarily from two datasets: the ONS death registration database, which provides date, place, and cause of death from February 2019 onwards; and primary care EHRs, which include recorded dates of death from 2009 onwards. Patients registered at GP practices managed by the software provider TPP were linked to ONS death registration database using NHS numbers via OpenSAFELY. To assign an NHS number, ONS mortality records are matched against the Personal Demographics Service (PDS). This is primarily an automated process, with manual tracing undertaken when an NHS number cannot be assigned automatically^14^. The primary care database and associated linkages available within the OpenSAFELY-TPP backend are refreshed every four weeks, incorporating newly available data and updates to existing records.

##### Box 1. Mortality data sources and recording processes

###### ONS death registration

In England, there is a statutory process for death certification and registration. Most deaths are certified by a medical practitioner using a Medical Certificate of Cause of Death (MCCD)^8^. The Medical Examiner then confirms the cause, and a statutory five-day period is allowed for registration with the Local Registration Service^8,15^. Registration records are transmitted electronically to the Office for National Statistics where quality assurance procedures are conducted, and International Classification of Diseases, Tenth Revision (ICD-10) codes are assigned to causes of death^8^. The ONS mortality register database is updated regularly with new and amended registrations^8^. The ONS death registration database in OpenSAFELY is updated monthly and includes information on the date, place, and cause of death.

We considered the ONS death registration database the gold standard for mortality information because it is based on statutory death registration and undergoes multiple validation procedures. However, deaths are not available immediately after occurrence. Following implementation of the statutory Medical Examiner system in September 2024, the median delay between death and registration was eight days, although registration times were longer for deaths certified by a coroner^8^. As a result, most recent deaths may not yet be available in ONS records at the time of data extraction. Deaths of usual residents of England and Wales that occur abroad, including Scotland and Northern Ireland, are not required to be registered, and the mortality dataset may include deaths registered in England and Wales regardless of residency status, including visitors and residents of other UK nations^8^.

###### Primary care EHRs

GP practices can record the death of a registered patient in the primary care EHR. In TPP SystmOne, clinical or administrative staff can use a dedicated system function to record the date of death when the practice becomes aware of a patient’s death; this process also generates a clinical code indicating death in the patient’s record. Death-related clinical codes may also be present in the EHR independently of this process if entered directly by practice staff.

Although a 2021 Data Provision Notice mandates timely updates of death notifications to the Personal Demographics Service (PDS) – the central national database of NHS patients in England, Wales, and the Isle of Man – GP practices are responsible only for recording community deaths^16^. However, there is no explicit statutory requirement to record all deaths in primary care EHRs. For deaths occurring in a hospital or when the GP is not the clinician issuing the MCCD, notification to primary care may not be immediate or systematic^16^. As a result, death-related variables in primary care EHRs may be incomplete, delayed, or inaccurate.

Separately, registration of a death by the General Register Office initiates an administrative process whereby the death is recorded on the patient’s PDS record, generating a deduction from the patient’s GP practice. Therefore, a patient may have a GP registration end date even if their death is not recorded in the primary care EHR.

### Study population and main variables

We included individuals with a recorded date of death in either the primary care EHRs or the ONS death registration database who were registered with a TPP practice on the day prior to death. For analyses assessing the timing of GP registration end relative to death, we additionally included individuals whose registration had ended up to 30 days before their date of death. We excluded individuals aged over 110 years due to the likelihood of data errors, and individuals with sex recorded as other than male or female due to disclosure risk (small numbers).

For each individual, we extracted the date of death from the ONS death registration dataset. From primary care EHRs, we extracted the date of death recorded in the dedicated date-of-death field. We also extracted the date associated with SNOMED CT codes indicating death. These codes were identified using the Primary Care Domain Reference Set Patient Deceased codelist, published and maintained by NHS England as a standardised definition of codes indicating death^17^. Finally, we extracted GP registration start and end dates from primary care EHRs to examine the temporal relationship between deregistration and death.

We extracted a set of demographic and context variables to support subgroup population analyses: sex, ethnicity and age bands (0–44, 45–64, 65–74, 75–84, and ≥85 years), patient’s registered practice (using an anonymous practice identifier), region, rural–urban classification of residence and Index of Multiple Deprivation (IMD) grouped by decile, based on the individual’s address at the reference date of death. The reference date was defined hierarchically as the ONS-registered date of death, the primary care EHR-recorded date of death, or the date of a death-related SNOMED code, in that order.

### Analysis

#### Primary care death records compared with ONS-registered deaths

We used the date of death from the ONS death registration dataset as the reference standard to assess the completeness and concordance of dates of death recorded in primary care EHRs. For each ONS-registered death, we classified those with a date of death also recorded in primary care EHRs by the number of days between the primary care and ONS death dates. Those without a primary care date of death were classified as ONS-only with or without a SNOMED CT code indicating death. Analyses were conducted by calendar year, in the overall population, and across population subgroups and general practices. As a secondary analysis, we calculated the difference in days between the date associated with the SNOMED CT code indicating death and the ONS date of death.

#### ONS mortality records’ dates of death and GP registration end dates

We calculated the difference between the GP registration end date and the date of death in ONS mortality records and described the distribution of registration ending before, on, and after the date of death. Individuals without a recorded registration end date were retained as a separate category. Individuals whose registration ended more than 30 days before death were excluded to focus the analysis on registration end dates occurring close to death, when deregistration may reflect processes around death Among the remaining population, we assessed the timing of registration end relative to death by calendar year, in the overall population, and across population subgroups. We also described the proportion of individuals with a missing registration end date among those with a date of death in ONS mortality records but no date of death recorded in primary care records.

#### Primary care death records without an ONS mortality record

We assessed the completeness of ONS mortality records compared with deaths recorded in primary care. Among deaths identified in either source in patients registered with a TPP practice, we calculated the proportion of deaths recorded in primary care without a corresponding ONS death registration, by calendar year, in the overall population, and across population subgroups. Deaths occurring close to the study end date were additionally examined by month to assess changes in the proportion of deaths without a corresponding ONS mortality record, potentially reflecting delays in death registration or in data availability.

#### Statistical Disclosure Control

All counts were rounded to the nearest five, and counts less than eight were suppressed.

## Results

Between 2020 and 2025, 1,324,995 deaths were recorded in primary care records among patients registered with TPP at the time of death, with a mean of 220,832 deaths per year. The highest annual number was recorded in 2020 (232,290 deaths). Over the same period, 1,409,275 deaths were identified through linked ONS mortality records among patients registered with TPP at the time of death, with a mean of 234,879 deaths per year. The highest annual number was recorded in 2020 (245,970 deaths).

### Primary care death records compared with ONS mortality records

Among 1,409,275 individuals with an ONS registered date of death between 2020 and 2025 who were registered with a TPP practice at the time of death, 7.5% (105,135/1,409,275) had no date of death recorded in primary care. This proportion increased from 6.5% in 2020 to 8.7% in 2025 (Figure 1 or <u>Supplementary Table 1</u>). Overall, 4.8% (67,590/1,409,275) had no date of death recorded in primary care but had a death-related SNOMED code, while 2.7% (37,545/1,409,275) had neither a date of death nor a death-related SNOMED code recorded in primary care.

**Figure 1:**
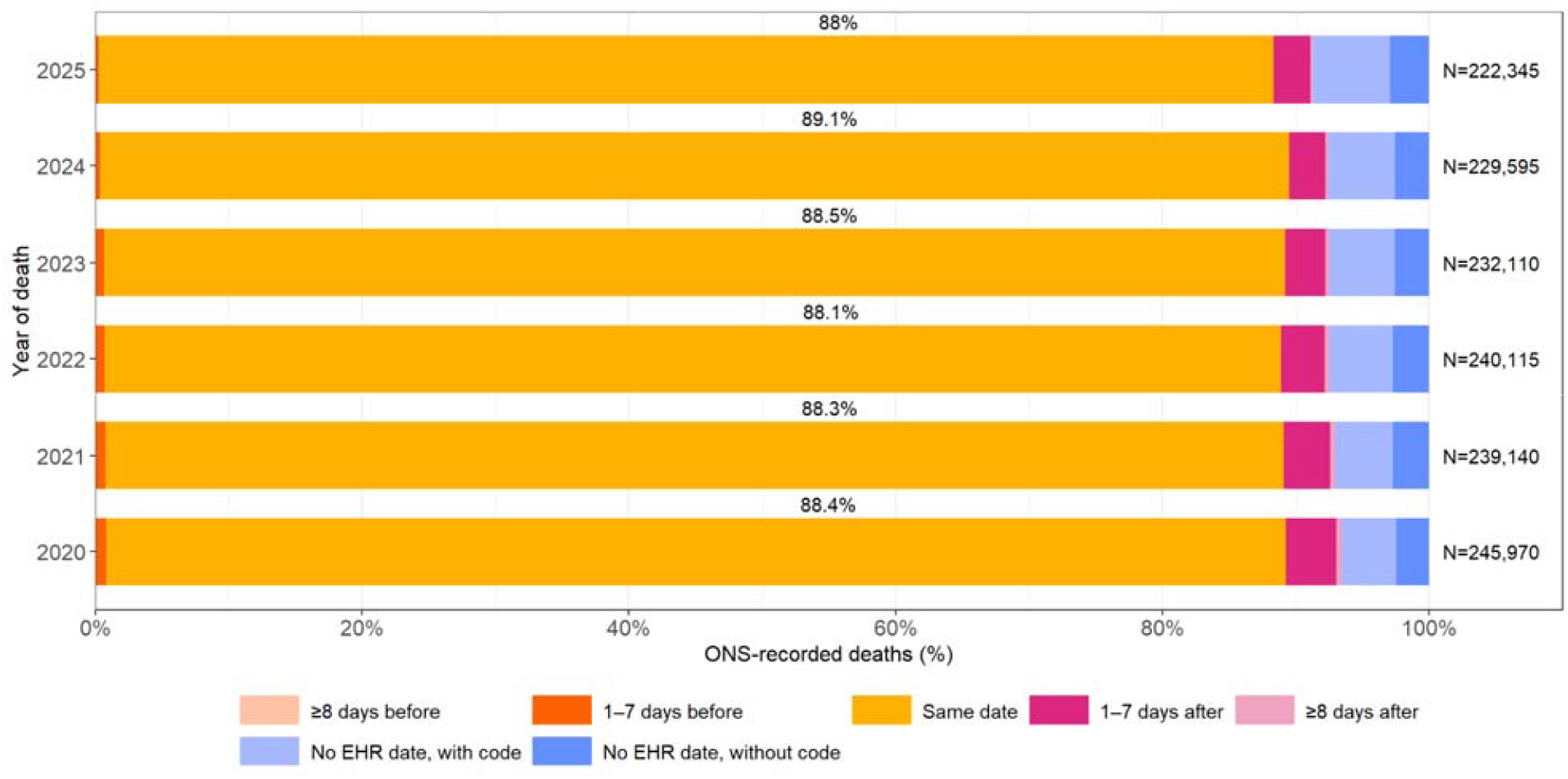
Primary care electronic health record (EHR) dates of death relative to Office fo National Statistics (ONS)-registered dates of death, by year. “Before” and “after” indicate that the date of death recorded in the primary care EHR was before or after the ONS-registered date of death, respectively. Among individuals without a date of death recorded in the primary care EHR, “with code” and “without code” indicate those who did or did not, respectively, have a death-related SNOMED CT code recorded. Percentages are calculated among all individuals with an ONS-registered date of death who were registered with a TPP practice at the time of death. The total number of individuals (N) is shown at the end of each bar for each year.

Across the study period, 88.4% (1,245,895/1,409,275) had the same date of death recorded in primary care as the ONS-registered date of death, while 4.1% (58,255/1,409,275) had a different date recorded in primary care. 3.5% (49,300/1,409,275) had a date of death recorded in primary care that was later than the ONS-registered date of death (3.2% (45,190/1,409,275) within 7 days). Variation across calendar years is shown in Figure 1 (or <u>Supplementary Table 1</u>). The timing of death-related SNOMED codes recorded in primary care relative to ONS-registered dates of death is shown in <u>Supplementary Figure 1</u>.

### Variation across population subgroups and practices

Across 2020–2025, the percentage of deaths without a same-day primary care date of death varied across demographic and geographic groups. Differences by sex and deprivation were relatively small, with similar percentages among males and females and no clear deprivation gradient. There was a consistent age-related pattern, with percentages decreasing as age increased. Differences by ethnicity were more apparent, with Black or Black British groups generally having higher percentages and White groups lower percentages, while estimates for other ethnic groups varied more between years.

Moderate variation was observed by rural/urban classification, with urban major conurbations generally among the highest, although there was no clear gradient and differences between categories varied over time. Regional variation was the most pronounced, with the West Midlands consistently having the highest, or among the highest percentages, and London also generally high. Detailed results by subgroup and year are presented in Figure 2 and <u>Supplementary Table 3</u>.

**Figure 2:**
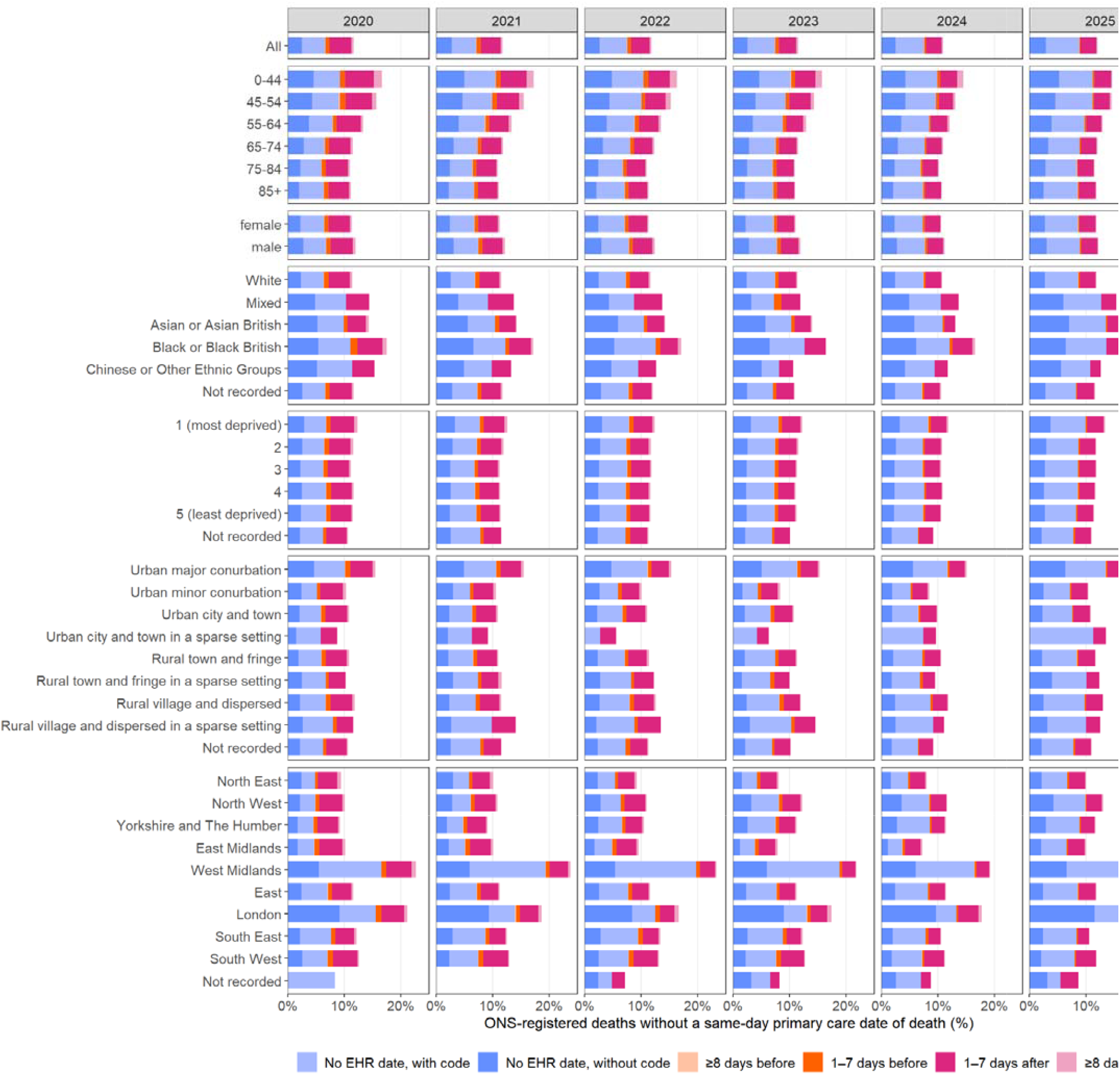
Percentage of ONS-registered deaths without a same-day primary care date of death, by year, population subgroup, and recording category. “Before” and “after” indicate that the date of death recorded in the primary care EHR was before or after the ONS-registered date of death, respectively. Among individuals without a date of death recorded in the primary care EHR, “with code” and “without code” indicate those who did or did not, respectively, have a death-related SNOMED CT code recorded. Percentages are calculated among all individual with an ONS-registered date of death within each year and population subgroup who were registered with a TPP practice at the time of death. All categories are mutually exclusive, and percentages use the same denominator: all individuals with an ONS-registered death within each year and population subgroup who were registered with a TPP practice at the time of death.

To examine variation across practices, we restricted the analysis to 2545 GP practices with more than 30 deaths among their registered patients in 2025 recorded in either ONS death registrations or primary care EHRs. In 30% of these practices, more than 12.5% of ONS-registered deaths had no corresponding date of death recorded in primary care; in 10% of practices, this proportion exceeded 30.8% (<u>Supplementary Figure 2</u>).

#### ONS-recorded dates of death and GP registration end dates

Among individuals with an ONS-registered date of death, after excluding those whose GP registration ended more than 28 days before death, the timing of GP registration end relative to the ONS-registered date of death varied across calendar years. GP registration increasingly ended on the ONS-registered date of death, rising from 8.6% (21,315/247,430) in 2020 to 84.6% (190,480/225,115) in 2025, with the largest increase between 2023 (9.2% (21,460/233,735)) and 2024 (65.9% (153,015/232,180)). Registration ending 1–28 days before the ONS-registered date of death remained uncommon but increased from 0.6% (1,465/247,430) in 2020 to 1.2% (2,770/225,115) in 2025, while registration ending after the ONS-registered date of death decreased from 90.8% (224,635/247,430) to 14.1% (31,670/225,115) over the same period. Among registrations ending after the ONS-registered date of death in 2025, 81.4% (25,790/31,670) ended within 7 days (Figure 3). Variation across population subgroups and calendar years is shown in <u>Supplementary Figure 3</u>.

**Figure 3:**
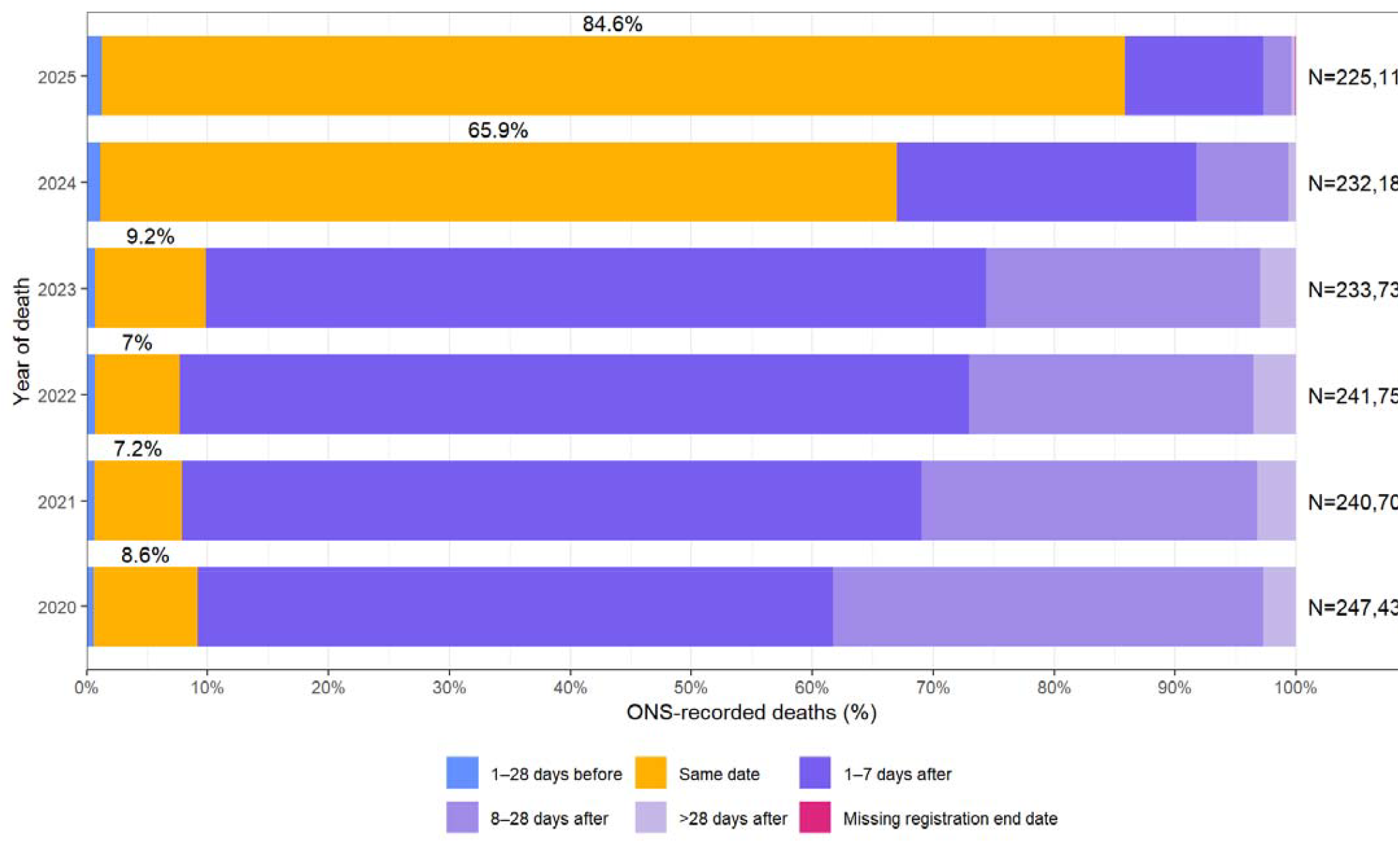
Timing of GP registration end relative to Office for National Statistics (ONS)- registered dates of death, by year. “Before” and “after” indicate that the GP registration end date occurred before or after the ONS-registered date of death, respectively. Percentages are calculated among all individuals with an ONS-registered date of death, after excluding those whose GP registration ended more than 28 days before death. The number of individuals with a missing GP registration end date was 15 in 2020, 20 in 2021, 20 in 2022, 25 in 2023, 65 in 2024, and 190 in 2025.

### Primary care death records without an ONS mortality record

Among all individuals with a date of death recorded in the primary care EHR between 2020 and 2025, 1.6% (20,865/1,324,995) lacked a corresponding ONS-registered date of death. The proportion increased from 1% in 2020 to 3.4% in 2025, with the largest increase occurring between 2024 (1.6%) and 2025. The proportion was higher for deaths occurring closer to the end of the study period, exceeding 4% from September 2025 and reaching 7.3% in February 2026 (Figure 4).

**Figure 4:**
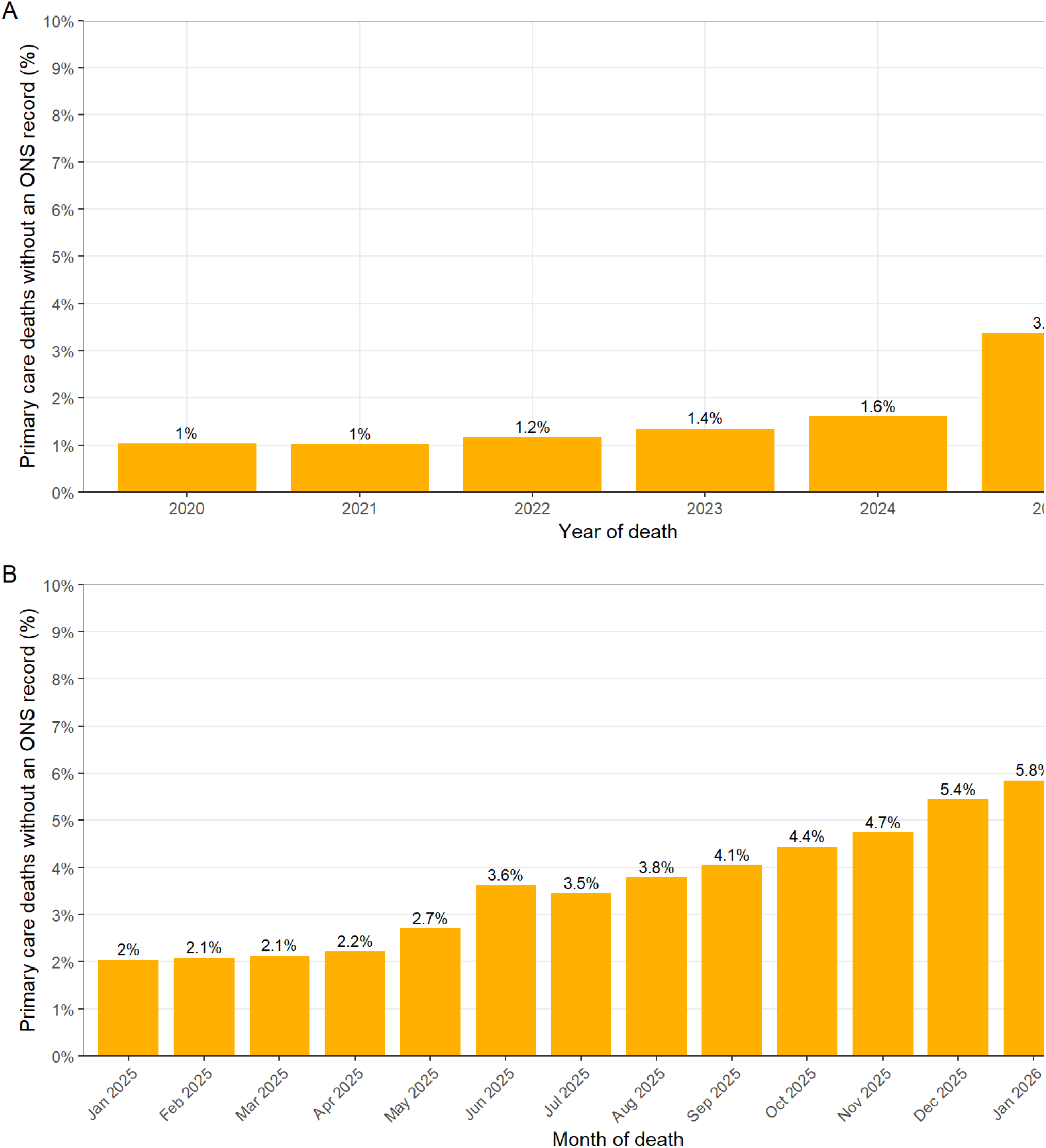
Dates of death recorded in the primary care electronic health record (EHR) without a corresponding Office for National Statistics (ONS)-registered date of death over time. (A) Annual proportions from 2020 to 2025. (B) Monthly proportions were calculated for 2025–2026 to provide greater temporal resolution for the most recent period. Percentages are calculated among all individuals with a date of death recorded in the primary care EHR.

Across 2020–2025, the percentage of primary care deaths without a corresponding ONS death registration varied across demographic and geographic groups. Differences by sex and deprivation were relatively small, with similar percentages among males and females and no clear deprivation gradient. Differences by age were more pronounced, particularly in 2025, when percentages were highest among those aged 0–44 years (25.8% (1,535/5,955)) and 45–54 years (11.9% (835/7,035)) and declined with increasing age. Differences by ethnicity were evident, with the White group consistently having the lowest percentages, while percentages were higher across several other ethnic groups, with the highest values observed in 2025 among Mixed, Asian or Asian British, Black or Black British, and Chinese or Other Ethnic Groups, reaching 12.2% (90/740), 10% (515/5,160), 9.5% (185/1,940) and 9% (95/1,055), respectively. Differences by rural/urban classification were relatively small, with no clear or consistent gradient across categories. Regional variation was more pronounced, with London generally having one of the highest percentages, although regional differences varied across years. Detailed results by subgroup and year are presented in Figure 5 and <u>Supplementary Table 5</u>.

**Figure 5:**
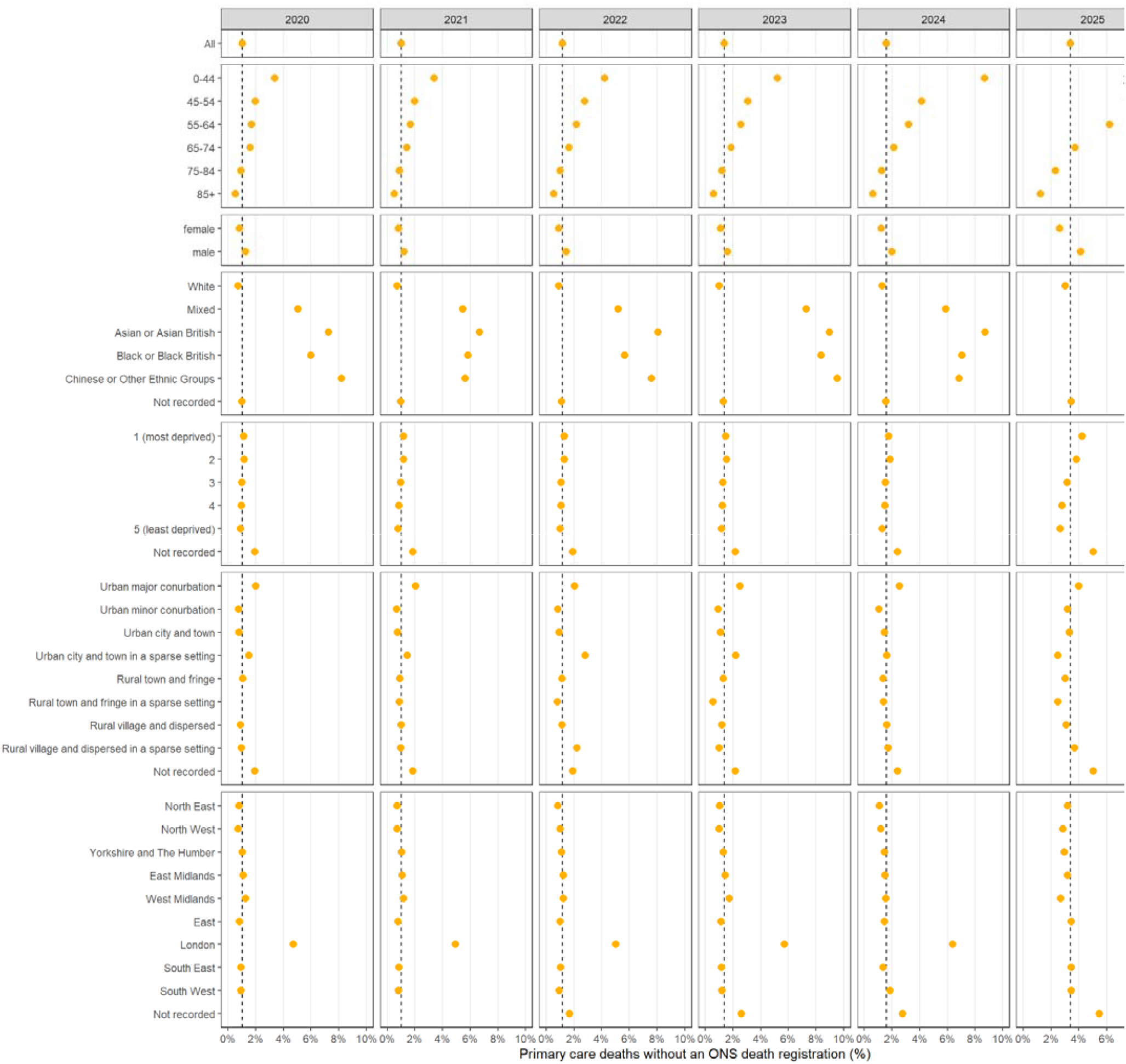
Dates of death recorded in the primary care electronic health record (EHR) without a corresponding Office for National Statistics (ONS) death registration, overall and by population subgroup and year. Percentages are calculated among all individuals with a date of death recorded in the primary care EHR within each subgroup and year. Points show subgroup-specific percentages, and dashed lines show the overall percentage for each year. Values above 10% are shown at the upper plotting boundary and labelled with their observed percentage (□).

## Discussion

#### Box 2. Recommendations for the definition and use of date of death in OpenSAFELY-TPP where ONS mortality data linkage is available

When ONS mortality data linkage is available:

- Use ONS death registrations as the primary source of mortality information.
- When analysing recent periods, assess the completeness of ONS mortality data by comparing it with primary care death dates to inform the choice of study end date.
- Assess whether mortality ascertainment differs across population groups, particularly by age, ethnicity, and geography.
- Where incomplete ONS mortality data may affect the analysis, consider complementing ONS data with information from primary care EHRs, including recorded dates of death and death-related SNOMED CT codes.

When only primary care EHR data are available:

- Consider combining recorded dates of death with death-related SNOMED CT codes to improve mortality ascertainment. The following codelist can be used to identify death-related SNOMED CT codes: opencodelists.org/codelist/nhsd-primary-care-domain-refsets/death_cod
- Consider the potential for incomplete and differential ascertainment of mortality across population groups and general practices when using primary care EHR data alone.

In all cases, transparently document the sources and definitions used to ascertain death and any identified data quality limitations.

We assessed the completeness and concordance of death recording in primary care EHRs compared with ONS death registrations in England using OpenSAFELY-TPP. The vast majority of deaths were recorded in both sources and, among these, dates of death were highly concordant, with most recorded on the same date. However, a non-negligible proportion of ONS-registered deaths had no corresponding date of death in primary care EHRs. Deaths recorded only in primary care were uncommon, but their proportion increased gradually over the study period, with a more pronounced increase in the most recent months. Discordance varied across population subgroups. Younger people and those from Mixed, Asian or Asian British, Black or Black British, and Chinese or Other ethnic groups had higher proportions of deaths recorded only in ONS death registrations or only in primary care EHRs, while differences by sex and area-level deprivation were small. Deaths recorded only in ONS death registrations showed geographical variation, with higher proportions in London and the West Midlands, as well as variation across general practices.

GP registration generally ended on or shortly after the date of death. Same-day deregistration became increasingly common over time and was the predominant pattern in 2024 and 2025. Few individuals remained registered after death, while deregistration in the 30 days preceding death was uncommon but became more common over time.

A key strength of this study is its scale and population coverage. The OpenSAFELY-TPP platform includes approximately 25 million individuals registered across more than 2,500 general practices in England, allowing assessment of mortality recording across a large and diverse population^12^. Individual-level linkage between primary care EHRs and ONS death registrations allowed assessment of the completeness and concordance of death recording across the two sources, as well as the relationship between recorded dates of death and GP registration status. ONS mortality records also undergo PDS matching, improving the reliability of linkage using NHS numbers^14^.

This study also has several limitations. Although we identified discrepancies between primary care EHRs and ONS death registrations, determining their underlying causes was outside the scope of this study. In particular, we did not have access to the date of death registration, limiting our ability to distinguish registration delays from other explanations for discordance between the two sources. Our analyses were descriptive and did not aim to estimate independent associations, so differences observed across subgroups may partly reflect overlap between population characteristics. ONS death registrations were available from 2019 onwards, with complete calendar years available from 2020. Several measures to improve mortality recording were introduced during the COVID-19 pandemic, meaning that patterns observed during our study period may not be transportable to earlier periods. Finally, although we used ONS death registrations as the gold standard for mortality data in England, they may still be subject to registration and reporting delays^8^. Some observed discrepancies may reflect linkage errors rather than true differences in death recording between the two sources.

The number of deaths identified in our study was consistent with the scale of OpenSAFELY-TPP, which includes approximately 24 million patients across around 43% of general practices in England. Between 2020 and 2025, our study population included 1,409,275 individuals with an ONS-registered date of death and 1,324,995 with a date of death recorded in the primary care EHR, equivalent to 43.1% and 40.5%, respectively, of the 3,269,411 deaths registered by ONS in England^18^. However, these comparisons are indicative rather than direct estimates of coverage, as TPP practices are not a random sample of general practices in England.

Two studies using CPRD reported greater completeness of death recording in primary care EHRs and lower, although steadily improving, concordance between primary care and ONS dates of death over time^10,11^. However, their findings are not directly comparable with ours because of important differences in data sources, processing, and study periods. Through OpenSAFELY-TPP, we used primary care EHR data from practices using TPP SystmOne in England, whereas the CPRD studies used pre-processed data from a subset of practices using EMIS and Vision systems^9^. CPRD analyses also used a derived date of death (cprd_ddate), generated using an unpublished algorithm that incorporates multiple variables from the patient record, and restricted analyses to practices and patients meeting predefined data quality criteria (“up-to-standard”; UTS)^9^. In addition, those studies covered earlier periods (1998–2015)^10,11^, whereas our analyses focused on 2020–2026, a distinction that is particularly relevant given the improvements in mortality recording and date concordance reported over time.

Our findings support the use of ONS mortality registrations as the preferred source of mortality information for research in England when linkage is available^10,11^. However, mortality ascertainment for recent periods of follow-up requires particular consideration. The increasing proportion of dates of death recorded in primary care EHRs without a corresponding ONS registration in the months closest to data extraction may reflect delays between death occurrence, registration, and availability in linked research datasets^13,15^. For studies using recent mortality data, examining temporal patterns and comparing ONS registrations with deaths recorded in primary care EHRs (through dates of death or death-related SNOMED CT codes) can inform assessment of the impact of incomplete recent data, the choice of study end date, and whether complementary primary care mortality data are needed. Incorporating the dates on which each death was registered and became available in the research dataset would enable a more direct assessment of these delays.

The proportions of dates of death recorded only in the ONS register or only in primary care EHRs varied across the population, particularly by age, ethnicity, geography, and general practice. Differential completeness may lead to underrepresentation of deaths and causes of death among some population groups in mortality research, potentially affecting the characterisation of mortality patterns and comparisons between groups. Potential explanations include linkage errors arising from difficulties assigning or validating NHS numbers through PDS matching, duplicate NHS numbers, or inconsistencies in the transfer of information between systems^20^; delays in death registration (including those associated with coroner involvement)^23^; deaths occurring outside England and Wales^8^; and variation in recording and administrative processes across general practices^11^; these mechanisms require further investigation.

Further research is needed to improve the identification and validation of death-related information in research using primary care EHRs linked to ONS mortality data. This could include evaluating algorithms that combine ONS-registered dates of death, primary care dates of death, death-related SNOMED CT codes, hospital records, and GP registration information. Further investigation of missing or delayed death recording across population groups, including more detailed disaggregation by age and ethnicity, could help quantify the potential for differential misclassification and bias. Analyses by place and cause of death could further help assess the potential impact of incomplete mortality ascertainment on studies of specific diseases and provide insight into possible mechanisms underlying observed discrepancies. Practice-level variation should also be investigated to determine whether observed discrepancies reflect systematic differences in recording or administrative processes across general practices. Finally, the increasing alignment of GP registration end dates with ONS-registered dates of death from 2024 onwards is encouraging, although the marked change from previous years requires further investigation. Assessing whether mortality data are fit for purpose, transparently documenting identified data quality issues, and clearly reporting the analytical approaches used to address them can strengthen the interpretability and reproducibility of research findings.

## Conclusion

Death recording in primary care EHRs showed high completeness and date concordance with ONS death registrations. However, non-negligible discrepancies remained, particularly for recent deaths and in some population groups, highlighting the potential for incomplete or differential mortality ascertainment. These findings highlight the importance of considering data recency when defining study periods and population-specific completeness when using linked mortality data for research.

### Patient and Public Involvement and Engagement (PPIE)

OpenSAFELY has involved patients and the public in various ways: we developed a public website that provides a detailed description of the platform in language suitable for a lay audience (https://opensafely.org); we have participated in two citizen juries exploring public trust in OpenSAFELY; we have co-developed an explainer video (https://www.opensafely.org/about/); we have patient representation who are experts by experience on our OpenSAFELY Oversight Board; we have partnered with Understanding Patient Data to produce lay explainers on the importance of large datasets for research; we have presented at various online public engagement events to key communities (e.g., Healthcare Excellence Through Technology; Faculty of Clinical Informatics annual conference; NHS Assembly; HDRUK symposium); and more. To ensure the patient voice is represented, we are working closely to decide on language choices with appropriate medical research charities (e.g., Association of Medical Research Charities).

## Data Availability Statement

All data and analysis code related to this study are available on GitHub (https://github.com/opensafely/death-report/).

## AI Disclosure Statement

We used GPT-5.6 and Grammarly v.1.2.291.1949 for language polishing, and GPT-5.6 to support data analysis coding tasks. All content was reviewed by the authors.

## Ethics Statement

This study was approved by the Health Research Authority (REC reference 20/LO/0651) and by the LSHTM Ethics Board (reference 21863).

