## Supplementary tables and figures for "Accuracy and completeness of death recording in primary care electronic health records compared with national death registrations in England using OpenSAFELY-TPP"

### Supplemental Materials

#### Supplementary tables

| Year of death | Primary care recording relative to ONS date of death | | | | | | |
| --- | --- | --- | --- | --- | --- | --- | --- |
|  | ≥8 days before | 1–7 days before | Same date | 1–7 days after | ≥8 days after | No EHR date, with code | No EHR date, without code |
| **2020 (N=245,970)** | 195 (<0.1%) | 1,900 (0.8%) | 217,400 (88.4%) | 9,390 (3.8%) | 980 (0.4%) | 9,990 (4.1%) | 6,110 (2.5%) |
| **2021 (N=239,140)** | 220 (<0.1%) | 1,670 (0.7%) | 211,170 (88.3%) | 8,335 (3.5%) | 805 (0.3%) | 10,380 (4.3%) | 6,565 (2.7%) |
| **2022 (N=240,115)** | 180 (<0.1%) | 1,645 (0.7%) | 211,630 (88.1%) | 7,910 (3.3%) | 790 (0.3%) | 11,490 (4.8%) | 6,475 (2.7%) |
| **2023 (N=232,110)** | 160 (<0.1%) | 1,440 (0.6%) | 205,415 (88.5%) | 7,075 (3%) | 660 (0.3%) | 11,355 (4.9%) | 6,005 (2.6%) |
| **2024 (N=229,595)** | 110 (<0.1%) | 820 (0.4%) | 204,540 (89.1%) | 6,285 (2.7%) | 485 (0.2%) | 11,455 (5%) | 5,905 (2.6%) |
| **2025 (N=222,345)** | 85 (<0.1%) | 530 (0.2%) | 195,740 (88%) | 6,195 (2.8%) | 390 (0.2%) | 12,920 (5.8%) | 6,485 (2.9%) |

Supp Table 1: **Primary care electronic health record (EHR) dates of death relative to Office for National Statistics (ONS)-registered dates of death, by year.** “Before” and “after” indicate that the date of death recorded in the primary care EHR was before or after the ONS-registered date of death, respectively. Among individuals without a date of death recorded in the primary care EHR, “with code” and “without code” indicate those who did or did not, respectively, have a death-related SNOMED CT code recorded. Percentages are calculated among all individuals with an ONS-registered date of death who were registered with a TPP practice at the time of death.

| Year of death | ONS deaths, N | Primary care date of death | | Primary care date of death or SNOMED code | |
| --- | --- | --- | --- | --- | --- |
|  |  | Recorded, n (%) | No date, n (%) | Recorded, n (%) | Neither, n (%) |
| **2020** | 245,970 | 229,870 (93.5%) | 16,100 (6.5%) | 239,860 (97.5%) | 6,110 (2.5%) |
| **2021** | 239,140 | 222,195 (92.9%) | 16,945 (7.1%) | 232,575 (97.3%) | 6,565 (2.7%) |
| **2022** | 240,115 | 222,150 (92.5%) | 17,965 (7.5%) | 233,640 (97.3%) | 6,475 (2.7%) |
| **2023** | 232,110 | 214,750 (92.5%) | 17,360 (7.5%) | 226,105 (97.4%) | 6,005 (2.6%) |
| **2024** | 229,595 | 212,235 (92.4%) | 17,360 (7.6%) | 223,690 (97.4%) | 5,905 (2.6%) |
| **2025** | 222,345 | 202,940 (91.3%) | 19,405 (8.7%) | 215,860 (97.1%) | 6,485 (2.9%) |

Supp Table 2: **Completeness of death recording in primary care electronic health records (EHRs), using dates of death alone or dates of death and death-related SNOMED CT codes.** Percentages are calculated among all individuals with an Office for National Statistics (ONS)-registered date of death who were registered with a TPP practice at the time of death. NA indicates suppressed values where the count was <8 in accordance with statistical disclosure control requirements.

| Population subgroup | 2020 | | 2021 | | 2022 | | 2023 | | 2024 | | 2025 | |
| --- | --- | --- | --- | --- | --- | --- | --- | --- | --- | --- | --- | --- |
|  | With code | Without code | With code | Without code | With code | Without code | With code | Without code | With code | Without code | With code | Without code |
| **Region** | | | | | | | | | | | | |
| North East | 2.4% (335/13,745) | 2.4% (330/13,745) | 2.9% (355/12,450) | 3.0% (370/12,450) | 3.1% (395/12,915) | 2.3% (300/12,915) | 2.7% (325/12,095) | 1.6% (190/12,095) | 3.1% (370/12,055) | 1.7% (205/12,055) | 4.5% (540/11,875) | 2.1% (250/11,875) |
| North West | 2.7% (705/26,440) | 2.1% (560/26,440) | 3.3% (810/24,740) | 2.8% (690/24,740) | 3.6% (905/25,120) | 2.8% (705/25,120) | 4.9% (1,200/24,515) | 3.2% (795/24,515) | 4.8% (1,160/24,190) | 3.7% (885/24,190) | 5.6% (1,310/23,315) | 4.2% (980/23,315) |
| Yorkshire and The Humber | 2.7% (990/36,650) | 1.7% (640/36,650) | 2.9% (985/34,390) | 1.9% (655/34,390) | 4.2% (1,465/35,285) | 2.4% (860/35,285) | 4.9% (1,690/34,145) | 2.6% (895/34,145) | 5.8% (1,970/33,845) | 2.8% (935/33,845) | 5.9% (1,940/32,680) | 2.9% (935/32,680) |
| East Midlands | 2.9% (1,285/43,870) | 1.7% (755/43,870) | 2.9% (1,250/42,915) | 2.2% (945/42,915) | 3.1% (1,340/42,910) | 1.7% (750/42,910) | 2.6% (1,075/41,830) | 1.3% (540/41,830) | 2.5% (1,045/41,545) | 1.2% (510/41,545) | 4.5% (1,790/40,115) | 2.1% (830/40,115) |
| West Midlands | 11.1% (1,105/9,915) | 5.4% (540/9,915) | 13.4% (1,240/9,260) | 5.9% (550/9,260) | 14.3% (1,285/8,955) | 5.4% (485/8,955) | 12.8% (1,120/8,745) | 6.0% (525/8,745) | 10.3% (865/8,365) | 6.1% (510/8,365) | 13.0% (1,055/8,085) | 6.6% (530/8,085) |
| East | 4.6% (2,500/54,050) | 2.4% (1,285/54,050) | 4.7% (2,555/54,130) | 2.4% (1,300/54,130) | 5.2% (2,720/52,815) | 2.6% (1,355/52,815) | 5.3% (2,690/50,670) | 2.4% (1,200/50,670) | 5.8% (2,940/50,840) | 2.5% (1,255/50,840) | 6.1% (3,020/49,375) | 2.4% (1,170/49,375) |
| London | 6.4% (490/7,655) | 9.1% (700/7,655) | 4.6% (340/7,375) | 9.3% (685/7,375) | 4.1% (275/6,675) | 8.4% (560/6,675) | 3.9% (255/6,460) | 9.1% (585/6,460) | 3.6% (220/6,090) | 9.7% (590/6,090) | 4.9% (290/5,915) | 11.5% (680/5,915) |
| South East | 5.5% (925/16,765) | 2.1% (360/16,765) | 5.8% (975/16,880) | 2.9% (490/16,880) | 6.6% (1,095/16,575) | 2.9% (475/16,575) | 6.2% (1,000/16,115) | 2.6% (420/16,115) | 5.8% (925/15,965) | 2.0% (325/15,965) | 5.9% (915/15,490) | 2.4% (365/15,490) |
| South West | 4.5% (1,650/36,755) | 2.5% (935/36,755) | 5.1% (1,865/36,875) | 2.4% (870/36,875) | 5.2% (1,995/38,250) | 2.5% (960/38,250) | 5.3% (1,970/36,930) | 2.3% (840/36,930) | 5.3% (1,930/36,130) | 1.9% (675/36,130) | 5.9% (2,045/34,855) | 2.1% (730/34,855) |
| Not recorded | 8.3% (10/120) | NA% (NA/120) | NA% (NA/125) | NA% (NA/125) | 2.4% (15/625) | 2.4% (15/625) | 3.3% (20/605) | 3.3% (20/605) | 4.4% (25/570) | 2.6% (15/570) | 2.3% (15/640) | 3.1% (20/640) |
| **Rural/urban** | | | | | | | | | | | | |
| Urban major conurbation | 5.4% (2,170/40,005) | 4.6% (1,850/40,005) | 5.7% (2,125/37,500) | 4.9% (1,830/37,500) | 6.3% (2,310/36,430) | 4.8% (1,740/36,430) | 6.2% (2,200/35,295) | 5.1% (1,815/35,295) | 6.1% (2,120/34,785) | 5.6% (1,945/34,785) | 7.1% (2,375/33,350) | 6.3% (2,105/33,350) |
| Urban minor conurbation | 2.7% (475/17,330) | 2.4% (415/17,330) | 3.0% (485/16,220) | 3.0% (480/16,220) | 3.3% (535/16,410) | 2.7% (435/16,410) | 2.7% (430/15,800) | 1.7% (270/15,800) | 3.2% (500/15,450) | 1.9% (300/15,450) | 4.6% (695/15,065) | 2.5% (375/15,065) |
| Urban city and town | 3.7% (4,810/128,770) | 2.0% (2,630/128,770) | 4.0% (5,090/126,515) | 2.3% (2,905/126,515) | 4.4% (5,600/126,565) | 2.2% (2,810/126,565) | 4.5% (5,460/121,525) | 2.1% (2,590/121,525) | 4.6% (5,560/120,330) | 2.0% (2,360/120,330) | 5.2% (6,085/116,375) | 2.3% (2,660/116,375) |
| Urban city and town in a sparse setting | 4.3% (30/690) | 1.4% (10/690) | 4.2% (30/710) | 2.1% (15/710) | 2.8% (20/715) | NA% (NA/715) | 4.3% (30/705) | NA% (NA/705) | 7.5% (50/670) | NA% (NA/670) | 11.2% (75/670) | NA% (NA/670) |
| Rural town and fringe | 4.0% (1,225/30,920) | 1.9% (590/30,920) | 4.3% (1,290/30,235) | 2.2% (660/30,235) | 4.7% (1,460/30,760) | 2.4% (725/30,760) | 5.4% (1,610/29,890) | 2.1% (635/29,890) | 5.2% (1,525/29,485) | 2.1% (620/29,485) | 6.1% (1,760/28,800) | 2.2% (635/28,800) |
| Rural town and fringe in a sparse setting | 4.2% (85/2,005) | 2.5% (50/2,005) | 4.4% (80/1,815) | 3.0% (55/1,815) | 5.5% (110/2,000) | 2.8% (55/2,000) | 5.0% (95/1,900) | 1.6% (30/1,900) | 5.0% (95/1,890) | 1.9% (35/1,890) | 6.0% (105/1,745) | 4.0% (70/1,745) |
| Rural village and dispersed | 4.6% (975/21,330) | 2.1% (455/21,330) | 4.7% (980/21,035) | 2.3% (485/21,035) | 5.3% (1,155/21,685) | 2.7% (575/21,685) | 5.8% (1,210/21,015) | 2.5% (520/21,015) | 6.2% (1,285/20,650) | 2.5% (510/20,650) | 7.3% (1,425/19,625) | 2.4% (480/19,625) |
| Rural village and dispersed in a sparse setting | 5.3% (90/1,685) | 2.7% (45/1,685) | 7.2% (120/1,675) | 2.7% (45/1,675) | 6.7% (115/1,705) | 2.1% (35/1,705) | 7.3% (120/1,645) | 3.0% (50/1,645) | 6.7% (105/1,570) | 2.5% (40/1,570) | 6.9% (110/1,600) | 3.1% (50/1,600) |
| Not recorded | 4.0% (130/3,235) | 2.2% (70/3,235) | 5.2% (180/3,440) | 2.6% (90/3,440) | 4.9% (190/3,845) | 2.3% (90/3,845) | 4.7% (205/4,335) | 2.2% (95/4,335) | 4.6% (220/4,770) | 1.9% (90/4,770) | 5.6% (285/5,110) | 2.2% (110/5,110) |
| **IMD quintile** | | | | | | | | | | | | |
| 1 (most deprived) | 3.8% (1,910/50,075) | 2.9% (1,440/50,075) | 4.3% (2,070/47,895) | 3.3% (1,585/47,895) | 4.8% (2,265/47,330) | 3.1% (1,465/47,330) | 4.8% (2,160/45,030) | 3.2% (1,450/45,030) | 5.0% (2,235/44,470) | 3.3% (1,465/44,470) | 6.2% (2,635/42,615) | 3.7% (1,580/42,615) |
| 2 | 3.9% (1,895/49,085) | 2.5% (1,245/49,085) | 4.2% (2,000/47,320) | 2.9% (1,395/47,320) | 4.6% (2,145/46,820) | 2.8% (1,305/46,820) | 4.8% (2,175/44,900) | 2.6% (1,165/44,900) | 4.7% (2,105/44,435) | 2.6% (1,160/44,435) | 5.6% (2,415/42,825) | 3.0% (1,275/42,825) |
| 3 | 4.0% (2,105/53,120) | 2.2% (1,180/53,120) | 4.2% (2,215/52,270) | 2.5% (1,290/52,270) | 4.9% (2,580/52,475) | 2.6% (1,350/52,475) | 5.0% (2,540/51,065) | 2.4% (1,235/51,065) | 4.9% (2,455/49,920) | 2.4% (1,185/49,920) | 5.8% (2,800/48,295) | 2.7% (1,320/48,295) |
| 4 | 4.3% (2,060/48,115) | 2.5% (1,190/48,115) | 4.3% (2,000/46,890) | 2.5% (1,195/46,890) | 4.9% (2,320/47,725) | 2.4% (1,150/47,725) | 4.9% (2,260/45,860) | 2.4% (1,080/45,860) | 5.2% (2,370/45,430) | 2.4% (1,085/45,430) | 5.9% (2,625/44,155) | 2.5% (1,125/44,155) |
| 5 (least deprived) | 4.5% (1,890/42,340) | 2.3% (985/42,340) | 4.6% (1,910/41,325) | 2.4% (1,005/41,325) | 4.7% (1,990/41,920) | 2.6% (1,110/41,920) | 4.9% (2,020/40,910) | 2.4% (985/40,910) | 5.1% (2,070/40,570) | 2.3% (920/40,570) | 5.5% (2,160/39,345) | 2.7% (1,075/39,345) |
| Not recorded | 4.0% (130/3,235) | 2.2% (70/3,235) | 5.2% (180/3,440) | 2.6% (90/3,440) | 4.9% (190/3,845) | 2.3% (90/3,845) | 4.7% (205/4,340) | 2.2% (95/4,340) | 4.6% (220/4,770) | 1.9% (90/4,770) | 5.6% (285/5,110) | 2.2% (110/5,110) |
| **Ethnicity** | | | | | | | | | | | | |
| White | 4.0% (6,865/170,845) | 2.3% (3,940/170,845) | 4.3% (7,360/172,195) | 2.6% (4,415/172,195) | 4.8% (8,710/183,095) | 2.5% (4,580/183,095) | 5.0% (9,195/185,380) | 2.5% (4,560/185,380) | 5.0% (9,335/186,185) | 2.4% (4,535/186,185) | 5.8% (10,685/182,845) | 2.7% (5,020/182,845) |
| Mixed | 5.5% (40/730) | 4.8% (35/730) | 5.2% (40/765) | 3.9% (30/765) | 4.4% (35/800) | 4.4% (35/800) | 4.0% (30/755) | 3.3% (25/755) | 5.6% (45/805) | 5.0% (40/805) | 6.7% (50/750) | 6.0% (45/750) |
| Asian or Asian British | 4.7% (270/5,745) | 5.2% (300/5,745) | 4.8% (285/5,935) | 5.6% (330/5,935) | 4.6% (235/5,090) | 5.9% (300/5,090) | 4.6% (230/5,035) | 5.8% (290/5,035) | 5.0% (265/5,295) | 5.9% (310/5,295) | 6.5% (350/5,375) | 7.0% (375/5,375) |
| Black or Black British | 5.7% (120/2,120) | 5.4% (115/2,120) | 5.6% (120/2,125) | 6.6% (140/2,125) | 7.3% (140/1,905) | 5.2% (100/1,905) | 6.2% (125/2,005) | 6.5% (130/2,005) | 5.9% (120/2,020) | 6.2% (125/2,020) | 7.1% (145/2,035) | 6.4% (130/2,035) |
| Chinese or Other Ethnic Groups | 6.2% (55/880) | 5.1% (45/880) | 4.9% (50/1,020) | 4.9% (50/1,020) | 4.8% (45/945) | 4.8% (45/945) | 3.1% (30/980) | 5.1% (50/980) | 5.2% (55/1,060) | 4.2% (45/1,060) | 5.1% (55/1,075) | 5.6% (60/1,075) |
| Not recorded | 4.0% (2,645/65,645) | 2.6% (1,680/65,645) | 4.4% (2,520/57,105) | 2.8% (1,600/57,105) | 4.8% (2,325/48,280) | 2.9% (1,415/48,280) | 4.6% (1,745/37,955) | 2.5% (945/37,955) | 4.8% (1,635/34,235) | 2.5% (855/34,235) | 5.4% (1,630/30,270) | 2.8% (850/30,270) |
| **Sex** | | | | | | | | | | | | |
| female | 4.1% (5,035/121,455) | 2.2% (2,705/121,455) | 4.3% (5,105/117,990) | 2.4% (2,845/117,990) | 4.7% (5,625/118,935) | 2.4% (2,855/118,935) | 4.9% (5,590/114,455) | 2.3% (2,650/114,455) | 5.0% (5,695/113,395) | 2.3% (2,655/113,395) | 5.8% (6,410/109,920) | 2.7% (3,005/109,920) |
| male | 4.0% (4,955/124,515) | 2.7% (3,405/124,515) | 4.4% (5,275/121,150) | 3.1% (3,720/121,150) | 4.8% (5,865/121,180) | 3.0% (3,620/121,180) | 4.9% (5,770/117,655) | 2.9% (3,355/117,655) | 5.0% (5,760/116,200) | 2.8% (3,250/116,200) | 5.8% (6,510/112,425) | 3.1% (3,480/112,425) |
| **Age group** | | | | | | | | | | | | |
| 0-44 | 4.6% (310/6,750) | 4.6% (310/6,750) | 5.4% (395/7,270) | 5.0% (365/7,270) | 5.6% (395/7,080) | 4.8% (340/7,080) | 5.5% (375/6,805) | 4.7% (320/6,805) | 5.7% (360/6,355) | 4.3% (275/6,355) | 5.9% (295/4,975) | 5.2% (260/4,975) |
| 45-54 | 4.7% (435/9,340) | 4.3% (405/9,340) | 5.2% (495/9,510) | 4.6% (440/9,510) | 5.7% (505/8,895) | 4.4% (390/8,895) | 5.4% (450/8,360) | 4.0% (335/8,360) | 5.4% (440/8,100) | 4.3% (345/8,100) | 6.6% (460/6,975) | 4.5% (315/6,975) |
| 55-64 | 4.2% (795/18,895) | 3.7% (700/18,895) | 4.6% (930/20,045) | 4.0% (795/20,045) | 5.0% (950/19,170) | 3.9% (745/19,170) | 5.2% (985/18,825) | 3.5% (665/18,825) | 5.0% (920/18,480) | 3.5% (650/18,480) | 5.9% (1,030/17,540) | 3.9% (680/17,540) |
| 65-74 | 3.8% (1,485/39,490) | 2.8% (1,095/39,490) | 4.2% (1,640/39,355) | 3.1% (1,210/39,355) | 4.7% (1,780/37,625) | 3.3% (1,225/37,625) | 4.6% (1,655/35,735) | 2.9% (1,035/35,735) | 4.7% (1,665/35,090) | 2.9% (1,030/35,090) | 5.5% (1,830/33,190) | 3.3% (1,090/33,190) |
| 75-84 | 3.7% (2,625/70,770) | 2.2% (1,575/70,770) | 4.0% (2,765/68,695) | 2.4% (1,655/68,695) | 4.3% (3,040/70,185) | 2.4% (1,710/70,185) | 4.6% (3,135/68,475) | 2.5% (1,700/68,475) | 4.6% (3,155/68,655) | 2.4% (1,645/68,655) | 5.5% (3,730/67,835) | 2.8% (1,905/67,835) |
| 85+ | 4.3% (4,340/100,725) | 2.0% (2,030/100,725) | 4.4% (4,160/94,265) | 2.2% (2,100/94,265) | 5.0% (4,820/97,155) | 2.1% (2,065/97,155) | 5.1% (4,755/93,920) | 2.1% (1,955/93,920) | 5.3% (4,920/92,920) | 2.1% (1,955/92,920) | 6.1% (5,570/91,830) | 2.4% (2,240/91,830) |
| **Overall** | | | | | | | | | | | | |
| All | 4.1% (9,990/245,970) | 2.5% (6,110/245,970) | 4.3% (10,380/239,140) | 2.7% (6,565/239,140) | 4.8% (11,490/240,115) | 2.7% (6,475/240,115) | 4.9% (11,355/232,110) | 2.6% (6,005/232,110) | 5.0% (11,455/229,595) | 2.6% (5,905/229,595) | 5.8% (12,920/222,345) | 2.9% (6,485/222,345) |

Supp Table 3: **Office for National Statistics (ONS)-registered deaths without a date of death recorded in the primary care electronic health record (EHR), by year and population subgroup.** “With code” and “without code” indicate individuals who did or did not, respectively, have a death-related SNOMED CT code recorded in the primary care EHR. Values are percentages, with the numerator and denominator shown in parentheses (n/N). Percentages are calculated among all individuals with an ONS-registered date of death within each year and population subgroup who were registered with a TPP practice at the time of death.

| Year of death | ONS deaths, N | Missing GP registration end date | |
| --- | --- | --- | --- |
|  |  | ONS only, n (%) | ONS & Primary Care, n (%) |
| **2020** | 247,430 | NA (NA) | 15 (<0.1%) |
| **2021** | 240,700 | NA (NA) | 15 (<0.1%) |
| **2022** | 241,750 | NA (NA) | 15 (<0.1%) |
| **2023** | 233,735 | NA (NA) | 20 (<0.1%) |
| **2024** | 232,180 | 15 (<0.1%) | 50 (<0.1%) |
| **2025** | 225,115 | 40 (<0.1%) | 150 (<0.1%) |

Supp Table 4: **Missing GP practice registration end dates among individuals with an Office for National Statistics (ONS)-registered date of death, according to whether the death was also recorded in the primary care electronic health record (EHR).** Percentages are calculated among all individuals with an ONS-registered date of death included in the registration-end analysis, after excluding those whose GP practice registration ended more than 28 days before death.

| Population subgroup | Year of death | | | | | |
| --- | --- | --- | --- | --- | --- | --- |
|  | 2020 | 2021 | 2022 | 2023 | 2024 | 2025 |
| **Overall** | | | | | | |
| **All** | 1.0% (2,425/232,290) | 1.0% (2,290/224,485) | 1.2% (2,635/224,785) | 1.4% (2,945/217,690) | 1.6% (3,465/215,700) | 3.4% (7,105/210,045) |
| **Age group** | | | | | | |
| **0-44** | 3.4% (215/6,345) | 3.4% (230/6,745) | 4.2% (280/6,625) | 5.2% (335/6,440) | 8.7% (545/6,265) | 25.8% (1,535/5,955) |
| **45-54** | 2.0% (170/8,670) | 2.0% (175/8,750) | 2.8% (230/8,230) | 3.1% (240/7,820) | 4.1% (315/7,625) | 11.9% (835/7,035) |
| **55-64** | 1.7% (300/17,700) | 1.7% (315/18,635) | 2.2% (390/17,865) | 2.6% (455/17,630) | 3.2% (560/17,465) | 6.2% (1,045/16,870) |
| **65-74** | 1.6% (600/37,510) | 1.4% (530/37,035) | 1.7% (585/35,210) | 1.9% (635/33,680) | 2.1% (700/33,095) | 3.7% (1,165/31,430) |
| **75-84** | 0.9% (635/67,210) | 0.9% (585/64,865) | 1.0% (665/66,105) | 1.2% (765/64,405) | 1.2% (805/64,660) | 2.3% (1,465/63,665) |
| **85+** | 0.5% (500/94,855) | 0.5% (450/88,460) | 0.5% (485/90,755) | 0.6% (510/87,720) | 0.6% (540/86,590) | 1.2% (1,060/85,085) |
| **Sex** | | | | | | |
| **female** | 0.8% (950/114,665) | 0.8% (915/110,955) | 0.9% (995/111,450) | 1.1% (1,165/107,385) | 1.2% (1,300/106,345) | 2.6% (2,705/103,210) |
| **male** | 1.3% (1,475/117,625) | 1.2% (1,375/113,530) | 1.4% (1,640/113,335) | 1.6% (1,775/110,305) | 2.0% (2,165/109,355) | 4.1% (4,395/106,835) |
| **Ethnicity** | | | | | | |
| **White** | 0.7% (1,180/161,225) | 0.7% (1,175/161,595) | 0.9% (1,530/171,335) | 1.0% (1,725/173,350) | 1.3% (2,270/174,585) | 3.0% (5,230/172,370) |
| **Mixed** | 5.0% (35/695) | 5.5% (40/730) | 5.2% (40/770) | 7.3% (55/755) | 5.9% (45/765) | 12.2% (90/740) |
| **Asian or Asian British** | 7.3% (405/5,580) | 6.7% (380/5,700) | 8.1% (400/4,950) | 9.0% (445/4,955) | 8.7% (450/5,170) | 10.0% (515/5,160) |
| **Black or Black British** | 6.0% (120/2,005) | 5.8% (115/1,975) | 5.6% (100/1,770) | 8.4% (160/1,910) | 7.0% (135/1,915) | 9.5% (185/1,940) |
| **Chinese or Other Ethnic Groups** | 8.2% (70/855) | 5.6% (55/975) | 7.6% (70/920) | 9.5% (95/995) | 6.8% (70/1,025) | 9.0% (95/1,055) |
| **Not recorded** | 1.0% (610/61,935) | 1.0% (530/53,510) | 1.1% (495/45,040) | 1.3% (465/35,730) | 1.6% (500/32,245) | 3.5% (995/28,780) |
| **IMD quintile** | | | | | | |
| **1 (most deprived)** | 1.1% (530/47,250) | 1.2% (535/44,775) | 1.3% (585/44,180) | 1.5% (620/42,045) | 1.8% (735/41,510) | 4.2% (1,700/40,100) |
| **2** | 1.2% (535/46,485) | 1.2% (535/44,455) | 1.3% (580/43,950) | 1.5% (650/42,215) | 1.8% (775/41,945) | 3.8% (1,555/40,690) |
| **3** | 1.0% (510/50,340) | 1.0% (480/49,245) | 1.1% (525/49,070) | 1.3% (610/47,900) | 1.5% (720/46,995) | 3.2% (1,440/45,615) |
| **4** | 1.0% (435/45,300) | 0.9% (380/44,075) | 1.1% (480/44,735) | 1.2% (525/43,045) | 1.5% (635/42,610) | 2.8% (1,165/41,575) |
| **5 (least deprived)** | 0.9% (355/39,815) | 0.8% (300/38,705) | 1.0% (400/39,215) | 1.2% (450/38,360) | 1.3% (495/38,075) | 2.7% (990/37,100) |
| **Not recorded** | 1.9% (60/3,095) | 1.9% (60/3,230) | 1.9% (70/3,635) | 2.2% (90/4,130) | 2.4% (110/4,570) | 5.0% (250/4,965) |
| **Rural/urban** | | | | | | |
| **Urban major conurbation** | 2.0% (730/36,720) | 2.1% (705/34,255) | 2.1% (680/33,055) | 2.5% (800/32,080) | 2.5% (800/31,525) | 4.0% (1,205/30,075) |
| **Urban minor conurbation** | 0.8% (125/16,565) | 0.7% (105/15,355) | 0.8% (130/15,570) | 0.9% (140/15,240) | 1.0% (155/14,805) | 3.2% (460/14,455) |
| **Urban city and town** | 0.8% (975/122,310) | 0.8% (905/119,425) | 0.9% (1,130/119,285) | 1.1% (1,280/114,755) | 1.5% (1,655/114,070) | 3.3% (3,695/111,325) |
| **Urban city and town in a sparse setting** | 1.5% (10/660) | 1.5% (10/680) | 2.8% (20/710) | 2.2% (15/685) | 1.6% (10/620) | 2.5% (15/605) |
| **Rural town and fringe** | 1.1% (315/29,420) | 0.9% (265/28,550) | 1.1% (330/28,905) | 1.3% (360/28,005) | 1.4% (375/27,710) | 3.0% (820/27,220) |
| **Rural town and fringe in a sparse setting** | NA% (NA/1,875) | 0.9% (15/1,690) | 0.8% (15/1,850) | 0.6% (10/1,785) | 1.4% (25/1,785) | 2.5% (40/1,615) |
| **Rural village and dispersed** | 0.9% (180/20,080) | 1.0% (205/19,775) | 1.1% (230/20,190) | 1.2% (235/19,520) | 1.6% (310/19,165) | 3.1% (565/18,290) |
| **Rural village and dispersed in a sparse setting** | 1.0% (15/1,570) | 1.0% (15/1,525) | 2.2% (35/1,585) | 1.0% (15/1,490) | 1.7% (25/1,450) | 3.7% (55/1,495) |
| **Not recorded** | 1.9% (60/3,095) | 1.9% (60/3,225) | 1.9% (70/3,635) | 2.2% (90/4,130) | 2.4% (110/4,570) | 5.0% (250/4,960) |
| **Region** | | | | | | |
| **North East** | 0.8% (105/13,185) | 0.7% (85/11,810) | 0.9% (105/12,320) | 1.0% (120/11,695) | 1.1% (125/11,605) | 3.2% (365/11,450) |
| **North West** | 0.7% (185/25,360) | 0.7% (170/23,410) | 1.0% (240/23,750) | 1.0% (225/22,740) | 1.2% (265/22,410) | 2.9% (620/21,645) |
| **Yorkshire and The Humber** | 1.0% (365/35,390) | 1.1% (350/33,100) | 1.1% (375/33,330) | 1.3% (420/31,980) | 1.4% (455/31,395) | 2.9% (905/30,710) |
| **East Midlands** | 1.1% (460/42,290) | 1.1% (445/41,165) | 1.3% (520/41,340) | 1.4% (585/40,800) | 1.5% (610/40,600) | 3.2% (1,240/38,735) |
| **West Midlands** | 1.3% (105/8,375) | 1.2% (90/7,560) | 1.2% (90/7,270) | 1.7% (125/7,220) | 1.5% (110/7,100) | 2.7% (180/6,680) |
| **East** | 0.8% (415/50,680) | 0.8% (395/50,670) | 1.0% (495/49,235) | 1.1% (535/47,315) | 1.4% (685/47,325) | 3.5% (1,615/46,800) |
| **London** | 4.7% (320/6,780) | 4.9% (330/6,685) | 5.0% (310/6,150) | 5.7% (340/5,960) | 6.4% (360/5,640) | 8.8% (480/5,430) |
| **South East** | 0.9% (145/15,625) | 0.9% (135/15,550) | 1.1% (160/15,155) | 1.2% (175/14,875) | 1.3% (200/14,915) | 3.5% (510/14,715) |
| **South West** | 0.9% (325/34,500) | 0.8% (285/34,425) | 1.0% (340/35,630) | 1.2% (410/34,530) | 1.9% (640/34,165) | 3.5% (1,150/33,235) |
| **Not recorded** | NA | NA% (NA/115) | 1.7% (10/600) | 2.6% (15/575) | 2.8% (15/545) | 5.5% (35/640) |

Supp Table 5: **Dates of death recorded in the primary care electronic health record (EHR) without a corresponding Office for National Statistics (ONS) death registration, by year and population subgroup.** Values are n (%). Percentages are calculated among all individuals with a date of death recorded in the primary care EHR within each year and population subgroup.

#### Supplementary Figures

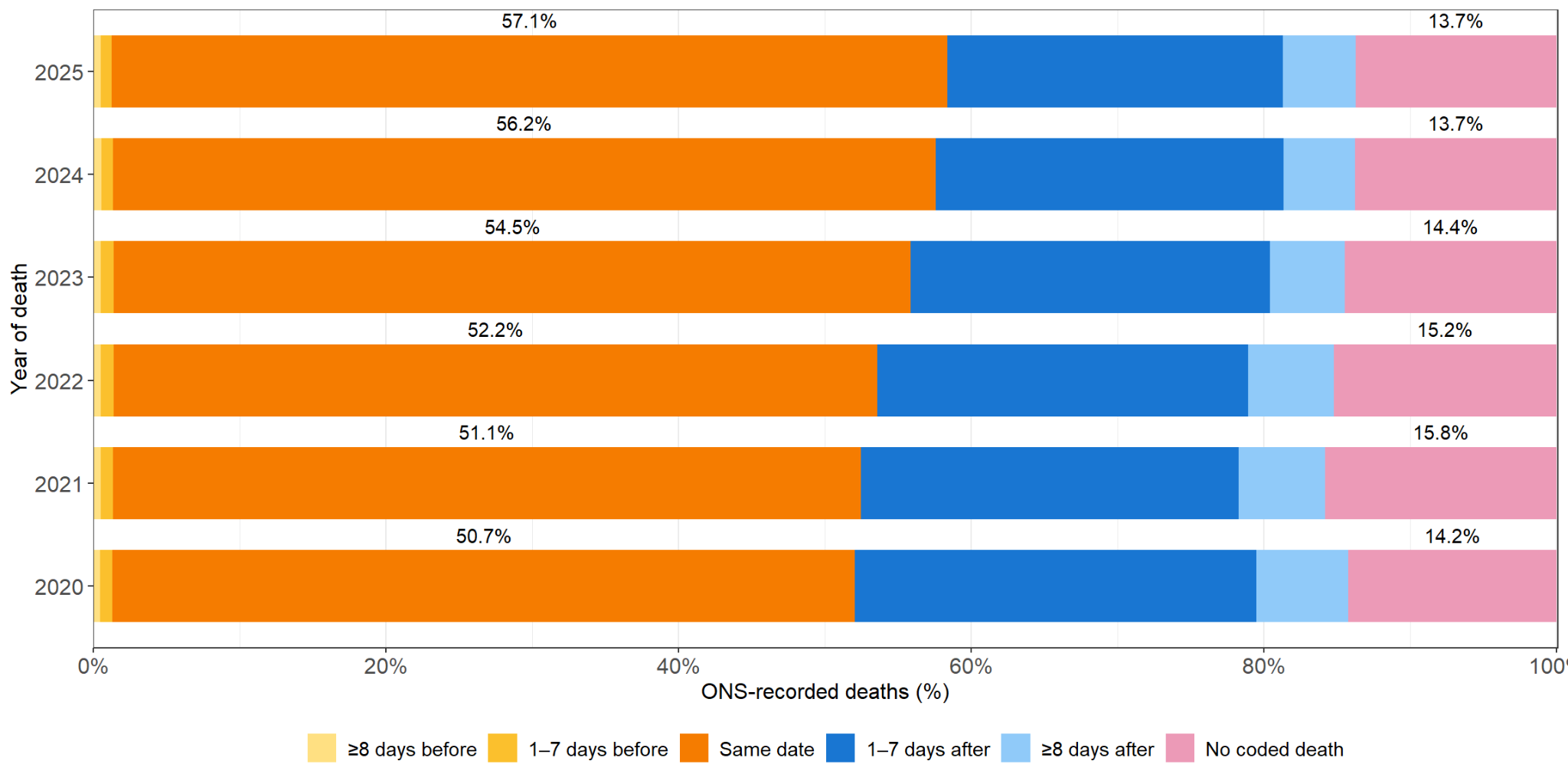

Supp Fig 1: **Timing of death-related SNOMED CT codes recorded in the primary care electronic health record (EHR) relative to Office for National Statistics (ONS)-registered dates of death, by year.** “Before” and “after” indicate that the death-related SNOMED CT code was recorded before or after the ONS-registered date of death, respectively. “No coded death” indicates individuals with an ONS-registered date of death who did not have a corresponding death-related SNOMED CT code recorded in the primary care EHR. Percentages are calculated among all individuals with an ONS-registered date of death represented in each year.

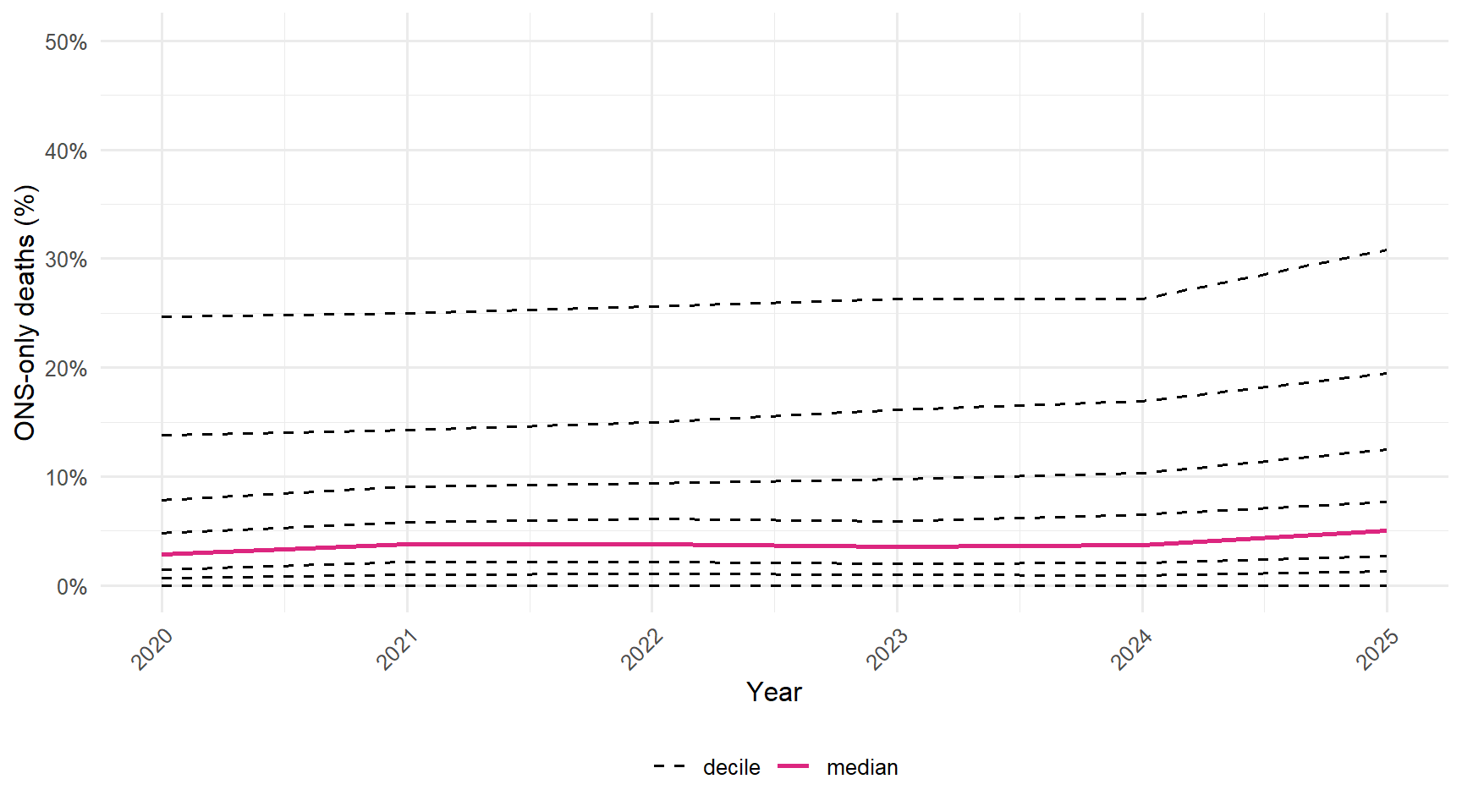

Supp Fig 2: **Distribution across general practices of the proportion of individuals with an Office for National Statistics (ONS)-registered date of death but no corresponding date of death recorded in the primary care electronic health record (EHR), 2020–2025.** Lines show practice-level percentiles of this proportion for each year.

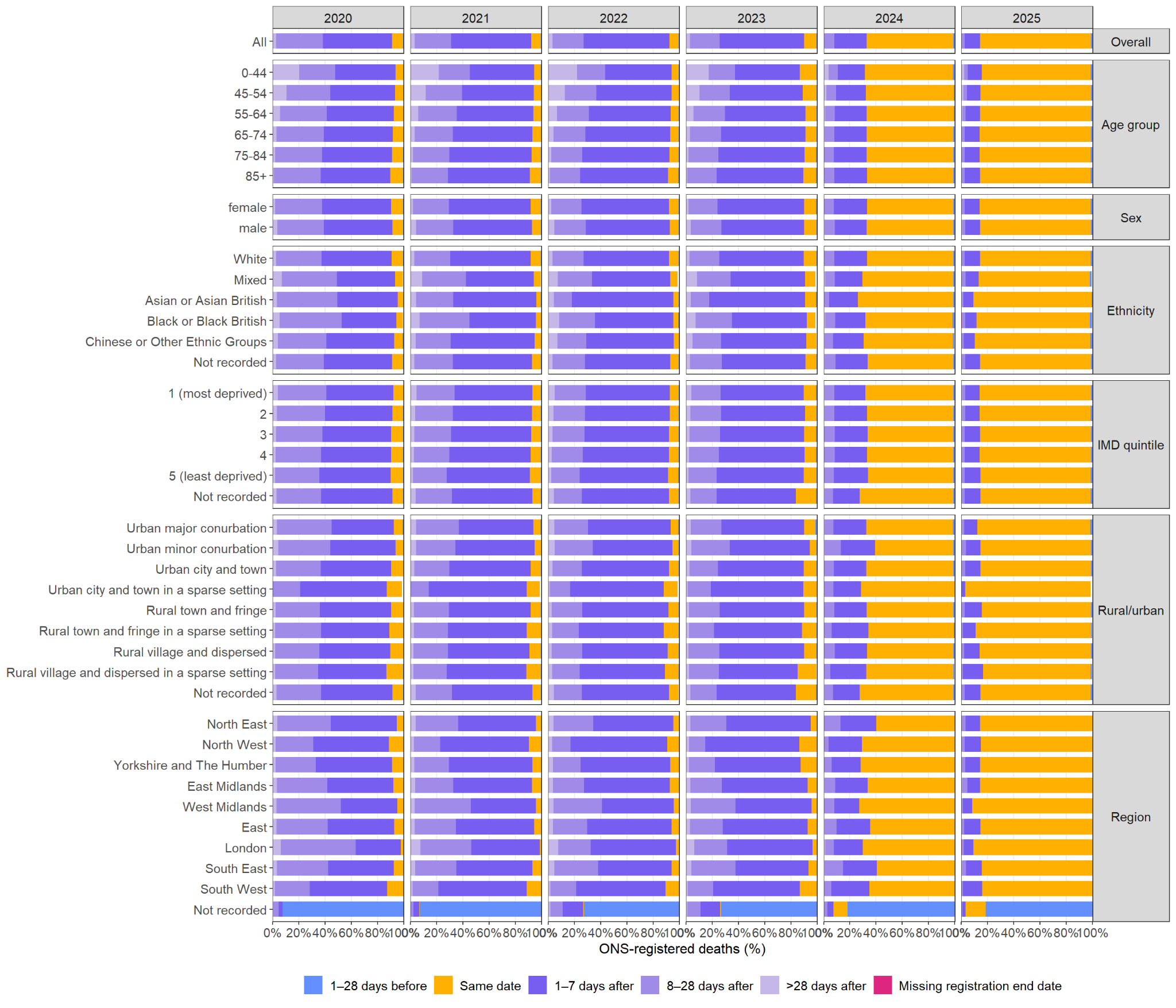

Supp Fig 3: **Timing of GP registration end relative to Office for National Statistics (ONS)-registered dates of death, overall and by population subgroup and year.** “Before” and “after” indicate that the GP registration end date occurred before or after the ONS-registered date of death, respectively. Percentages are calculated among individuals with an ONS-registered date of death within each subgroup and year, after excluding those whose GP registration ended more than 28 days before death. Individuals with no recorded GP registration end date remained in the denominator.
